# Prevalence and outcomes of co-infection and super-infection with SARS-CoV-2 and other pathogens: A Systematic Review and Meta-analysis

**DOI:** 10.1101/2020.10.27.20220566

**Authors:** Jackson Musuuza, Lauren Watson, Vishala Parmasad, Nathan Putman-Buehler, Leslie Christensen, Nasia Safdar

**Author notes:** Corresponding Author: (NS).

## Abstract

**Introduction:** The recovery of other respiratory viruses in patients with SARS-CoV-2 infection has been reported, either at the time of a SARS-CoV-2 infection diagnosis (co-infection) or subsequently (superinfection). However, data on the prevalence, microbiology and outcomes of co-infection and super infection are limited. The purpose of this study was to examine occurrence of respiratory co-infections and superinfections and their outcomes among patients with SARS-CoV-2 infection.

**Patients and Methods:** We searched literature databases for studies published from October 1, 2019, through June 11, 2020. We included studies that reported clinical features and outcomes of co-infection or super-infection of SARS-CoV-2 and other pathogens in hospitalized and non-hospitalized patients. We followed PRISMA guidelines and we registered the protocol with PROSPERO as: CRD42020189763.

**Results:** Of 1310 articles screened, 48 were included in the random effects meta-analysis. The pooled prevalence of co-infection was 12% (95% confidence interval (CI): 6%-18%, n=29, I^2^=98%) and that of super-infection was 14% (95% CI: 9%-21%, n=18, I^2^=97%). Pooled prevalence of pathogen type stratified by co- or super-infection: viral co-infections, 4% (95% CI: 2%-7%); viral super-infections, 2% (95% CI: 0%-7%); bacterial co-infections, 4% (95% CI: 1%-8%); bacterial super-infections, 6% (95% CI: 2%-11%); fungal co-infections, 4% (95% CI: 1%-8%); and fungal super-infections, 4% (95% CI: 0%-11%). Compared to those with co-infections, patients with super-infections had a higher prevalence of mechanical ventilation [21% (95% CI: 13%-31%) vs. 7% (95% CI: 2%-15%)] and greater average length of hospital stay [mean=12.5 days, standard deviation (SD) =5.3 vs. mean=10.2 days, SD= 6.7].

**Conclusions:** Our study showed that as many as 14% of patients with COVID-19 have super-infections and 12% have co-infections. Poor outcomes were associated with super-infections. Our findings have implications for diagnostic testing and therapeutics, particularly in the upcoming respiratory virus season in the Northern Hemisphere.

## Introduction

The coronavirus disease 2019 (COVID-19) pandemic is associated with high morbidity and mortality.(1, 2) Current evidence shows that transmission of severe acute respiratory syndrome coronavirus 2 (SARS-CoV-2), the causative agent of COVID-19, happens primarily through respiratory droplets (3, 4) from symptomatic, asymptomatic or pre-symptomatic individuals.(4, 5) Similar to other respiratory pathogens such as influenza, where approximately 25% of older patients get secondary bacterial infections,(6, 7) both super-infections and co-infections with SARS Cov-2 have been reported.(8-10) However, little data is available on the magnitude of co-infection and super-infections by viral, bacterial and fungal infections and associated clinical outcomes among patients infected with SARS-CoV-2.(8-10)

We define co-infection as the recovery of other respiratory pathogens in patients with SARS-CoV-2 infection at the time of a SARS-CoV-2 infection diagnosis and super-infection as the subsequent recovery of other respiratory pathogens during care for patients infected with SARS-CoV-2.

Two previous reviews have focused on this topic and examined the prevalence of bacterial and fungal co-infection or super-infection in SARS-CoV-2 infected patients.(11, 12) We extended this work by distinguishing between super-and co-infection because of the different implications of co-infections vs super-infections. For example, Garcia-Vidal et. al., showed that SARS-CoV-2 infected patients with superinfections had longer LOS and higher mortality while those with co-infections were had a higher frequency of admission to the ICU.(13) In addition, we examined the impact of co-infections vs. super-infections on clinical outcomes such as average length of hospital stay (LOS).

Diagnostic testing and therapeutic decision making may be affected by the presence of co-infection or super-infection with SARS-CoV-2 and other respiratory pathogens.

Therefore, we conducted a systematic review and meta-analysis to examine occurrence of and outcomes of respiratory co-infections and super-infections among SARS-CoV-2.

## Materials and methods

We conducted this systematic review in accordance with the Preferred Reporting in Systematic Reviews and Meta-Analyses (PRISMA) guidelines.(14) We registered this review with PROSPERO: CRD42020189763.(15) The protocol is available as a support document (S1. File).

### Data Sources and Searches

With the help of a health sciences librarian (LC), we searched PubMed, Scopus, Wiley, Cochrane Central Register of Controlled Trials, Web of Science Core Collection, and CINAHL Plus databases to identify English-language studies published from October 1, 2019 through June 11, 2020. We executed the search in PubMed and translated the keywords and controlled vocabulary for the other databases and additional articles were added from reference lists of pertinent articles. The following key words were used for the search: “coronavirus”,”coronavirus infections”, “HCoV”, “nCoV”, “Covid” “SARS”, “COVID-19”, “2019 nCoV”, “nCoV 19”, “SARS-CoV-2”, “SARS coronavirus2”, “2019 novel corona virus”, “Human”, “pneumonia”, “influenza”, “severe acute respiratory syndrome”, “co-infection”, “Super-infection”, “bacteria”, “fungus”, “concomitant”, “pneumovirinae”, “pneumovirus infections”, “respiratory syncytial viruses”, “metapneumovirus”, “influenza”, “human”, “respiratory virus”, “bacterial Infections”, “viral infection”, “fungal infection”, “upper respiratory”, “oxygen inhalation therapy”, “intensive care units”, “nursing homes”, “subacute care”, “skilled nursing”, “intermediate care”, “patient discharge”, “mortality”, “morbidity” and English filter. A complete description of our search strategy is available as supplementary material (S2. File).

### Study Selection

Citations were uploaded into Covidence®, an online systematic review software for the study selection process. Two authors (JSM and LW) independently screened titles and abstracts and read the full texts to assess if they met the inclusion criteria. The authors met and discussed any articles where there was conflict and decided to either include or exclude such articles. Inclusion criteria were randomized clinical trials (RCTs), quasi-experimental and observational human studies that reported clinical features and outcomes of co-infection or super-infection of SARS-CoV-2 (laboratory confirmed) and other pathogens—fungal, bacterial or other viruses in hospitalized and non-hospitalized patients. We excluded studies that did not report co-infection or super-infection, editorials, reviews, qualitative studies, those published in non-English language, articles where full texts were not available, and non-peer reviewed preprints.

### Data Extraction and risk of bias assessment

Three reviewers (JSM, LW, and VP) independently abstracted data from individual studies using a standardized template. We abstracted data on: study design/methodology, location and setting (intensive care unit (ICU), inpatient non-ICU, or outpatient, where applicable), study population, proportion of patients with co-infections, implicated pathogens, method of detection of co-infections and super-infections (laboratory verified or clinical features only), type of infection— bacterial, viral or fungal, outcomes of co-infected patients— death, ICU admission, mechanical ventilation, discharge disposition, length of hospital stay, or mild illness. Discrepancies were resolved by discussion between the three abstractors.

Risk of bias assessment was conducted by two authors (JSM and LW) independently. We used a tool developed in 2018 by Murad et al.(16) This tool examines four domains: selection, ascertainment, causality and reporting. The selection domain helps to assess whether participants included in a study are representative of the entire population from which they arise. Ascertainment assesses whether the exposure and outcome were adequately ascertained. Causality assesses the potential for alternative explanations and specifically for our study whether the follow-up was long enough for outcomes to occur. Reporting evaluates if a study described participants in sufficient detail to allow for replication of the findings. This tool consists of 8 items, but only five were applicable to our study.(16) When an item was present in a study, a score of 1 was assigned and 0 if the item was missing. We added the scores (minimum of 0 and a maximum of 5) and assigned the risk of bias as follows: low risk (5), medium risk (3-4), high risk (0-2). Details of the risk of bias assessment are provided as supporting information (S1 Table)

### Data Synthesis and Analysis

The primary outcome was the prevalence of co-infections or super-infections by viral, bacterial and fungal respiratory infections and SARS-CoV-2. We examined whether co-infection or super-infection was associated with an increased risk for the following patient outcomes: 1) mechanical ventilation, 2) admission to the ICU, and 3) mortality. We estimated the proportion of patients with co-infection or super-infection of viral, bacterial and fungal respiratory infections and SARS-CoV-2. We anticipated a high-level of heterogeneity given the novelty of COVID-19 and potential differences in testing and management of COVID-19 in the healthcare systems of the countries where the studies were conducted. We conducted all statistical analyses using Stata software, version 16.0 (Stata Corp. College Station, Texas). We used the “metan” and “metaprop” commands in Stata to estimate the pooled proportion of co-infection and super-infection and COVID-19 using a random effects model (DerSimonian Laird).(17, 18) We stabilized the variance using the Freeman-Tukey arcsine transformation methodology in order to correctly estimate extreme proportions (i.e., those close to 0% or 100%).(17) We assessed heterogeneity using the I^2^ statistic. Frequencies of outcome variables and study characteristics were estimated using descriptive statistics. For example, in studies where data on co-infecting or super-infecting pathogens was reported, we extracted and tallied the number of different pathogens reported. We calculated the proportion of pathogens using the number of pathogens as the numerator and the total number of pathogen type (bacteria, viruses and fungi) from all the studies as the denominator. We did not assess for publication bias because standard methods such as funnel plots and associated tests were developed for comparative studies and therefore do not produce reliable results for meta-analysis of proportions.(19, 20)

## RESULTS

Our search yielded 1840 records; we excluded 530 duplicates and screened 1310 articles. At the abstract and title review, we excluded 1231 articles leaving 79 articles for full text review. Of these, 48 articles met the inclusion criteria and were included in this meta-analysis. The most frequent reason for exclusion of studies at the full text review was the absence of super-infection or co-infection data (Figure 1).

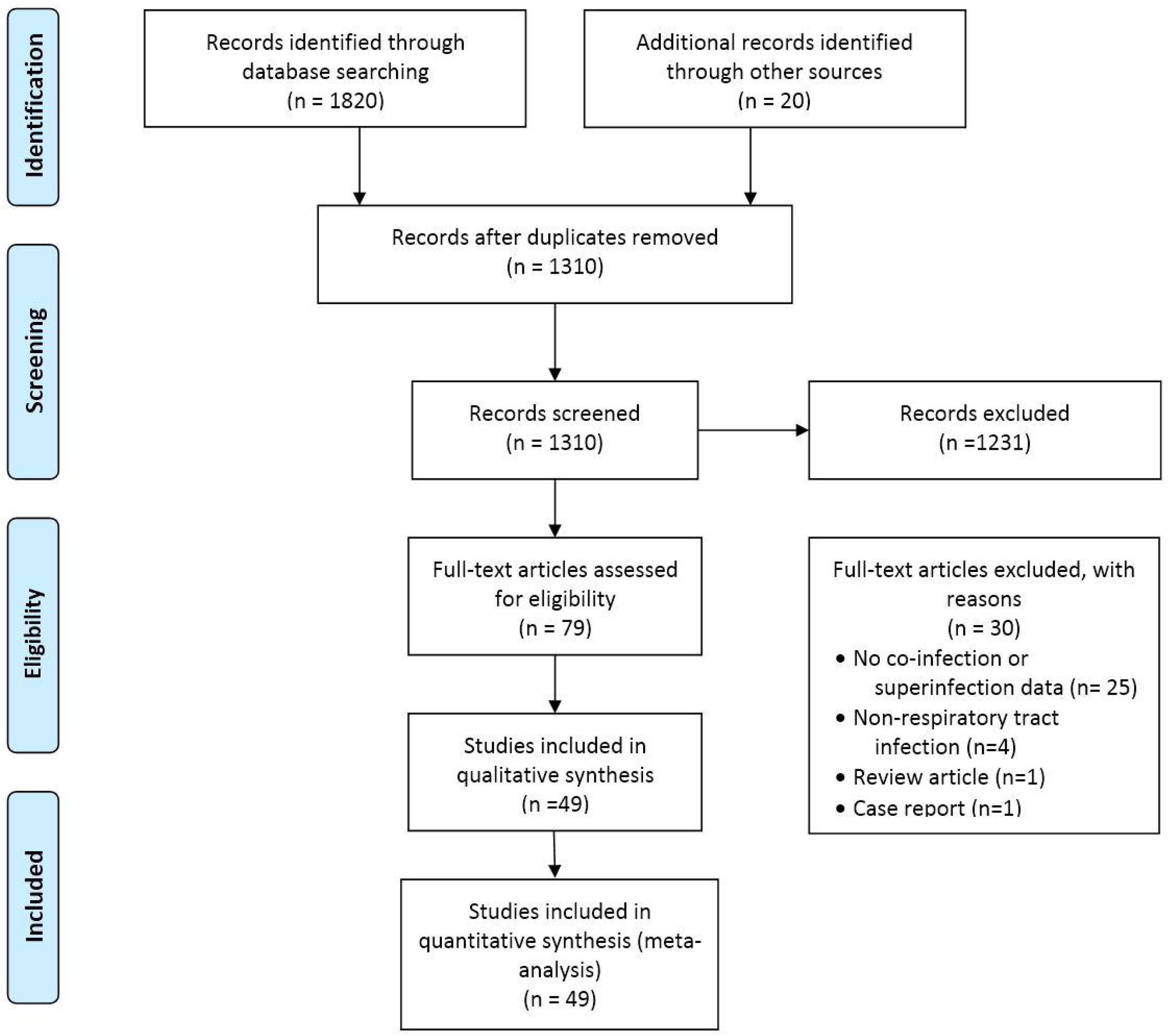

Most studies were case series (31/48). There were 14 retrospective cohort studies, 2 prospective cohorts and 1 RCT.(21) This RCT was a drug trial but also reported co-infection or super-infection data in patients with SARS-CoV-2. Sixty percent of the studies were conducted in China while 15% (7/48) were from the USA. Most of the studies were conducted in a mixed setting i.e., ICU and non-ICU setting (75% or 36/48), 92% (44/48) were conducted exclusively in hospitalized patients, and the majority were conducted among adults (71% or 34/48). Nineteen studies (48%) of the included studies in this review reported that patients included had super-infections, while co-infections were reported in 70% (30/43) of the studies. Viral co-infections in patients were reported in 74% (31/42) of the included studies, bacterial in 54% (26/48), fungal in 22% (8/37) of the studies. Seventy-three percent (27/37) of the studies reported at least one causative organism of the co-infection or super-infection (Table 1).

**Table 1.** Main characteristics of included studies.

| Study | Study design | Country | Setting | Number of patients | Age group of patients | Gender (% male) | ICU (%) | Patients who were ventilated n (%) | Patients who died n (%) | Viral co-infections n (%) | Bacterial co-infections n (%) | Fungal co-infections n (%) | Risk of bias |
| --- | --- | --- | --- | --- | --- | --- | --- | --- | --- | --- | --- | --- | --- |
| Arentz, 2020 (32) | Case series | USA | ICU<sup>a</sup> | 21 | Adults | 52 | 100 | 15 (71) | 11 (52) | 3 (14) | 1 (50) | 0 (0) | Medium |
| Barrasa, 2020 (33) | Case series | Spain | ICU | 48 | Adults | 56 | 100 | 45 (94) | 16 (33) | 0 (0) | 6 (13) | 0 (0) | Low |
| Campochiaro, 2020 (34) | Prospective cohort | Italy | ICU and non-ICU | 65 | Adults | 29 | 6 | 25 (38) | 16 (25) | 0 (0) | 1 (2) | 0 (0) | Low |
| Chen, 2020 (35) | Case series | China | ICU | 99 | Adults | 68 | 100 | 17 (17) | 11 (11) | 0 (0) | 1 (1) | 4 (4) | Medium |
| Cuadrado-Payán, 2020 (24) | Case series | Spain | ICU | 4 | Adults | 75 | 75 | 3 (75) | 0 (0) | 4 (100) | 0 (0) | 0 (0) | High |
| Ding, 2020 (36) | Case series | China | Non-ICU | 115 | Adults | NR<sup>b</sup> | 0 | 0 (0) | 0 (0) | 5 (4) | 0 (0) | 0 (0) | Medium |
| Dong, 2020 (22) | Case series | China | Non-ICU | 11 | Adults/children | 54 | 0 | 1 (9) | 0 (0) | 1 (9) | 0 (0) | 0 (0) | Medium |
| Du, 2020 (37) | Case series | China | ICU | 109 | Adults | 67.9 | 48.6 | 33 (30) | 109 (100) | 0 (0) | NR | NR | Low |
| Fan, 2020 (38) | Retrospective cohort | China | ICU and non-ICU | 50 | Adults | 83 | 54 | 23 (46) | 12 (24) | 0 (0) | 5 (10) | 5 (10) | Low |
| Feng, 2020 (39) | Case series | China | ICU and non-ICU | 476 | Adults | 56.9 | 26 | 70 (15) | 38 (8) | 0 (0) | 35 (7) | 0 (0) | Medium |
| Garazzino, 2020 (40) | Retrospective cohort | Italy | ICU and non-ICU | 168 | Children | 55.9 | 1.1 | 2 (1) | 0 (0) | 10 (6) | 1 (0.5) | 0 (0) | Low |
| Gayam, 2020 (41) | Case series | USA | ICU and non-ICU | 350 | Adults | 33 | NR | NR | NR | 0 (0) | 1 (0.3) | 0 (0) | Medium |
| Huang, 2020 (42) | Case series | China | ICU and non-ICU | 41 | Adults | 73 | 32 | 4 (10) | 6 (15) | 0 (0) | 1 (2) | 0 (0) | Medium |
| Kakuya, 2020 (43) | Case series | Japan | Non-ICU | 3 | Children | 100 | 0 (0) | 0 (0) | 0 (0) | 1 (33) | 0 (0) | 0 (0) | Low |
| Khodamordi, 2020 (44) | Case series | Iran | Non-ICU | 4 | Adults | 75 | 0 | 0 (0) | 0 (0) | 4 (100) | 0 (0) | 0 (0) | Medium |
| Kim, 2020 (45) | Retrospective cohort | USA | Non-ICU | 115 | Adults/children | 45 | 0 | 0 (0) | 0 (0) | 25 (22) | 0 (0) | 0 (0) | Low |
| Koehler, 2020 (46) | Case series | Germany | ICU | 19 | Adults | NR | 100 | NR | 3 (16) | 2 (11) | 0 (0) | 5 (26) | Medium |
| Lian, 2020 (47) | Retrospective cohort | China | ICU and non-ICU | 788 | Children/Adults | 52 | 3 | 18 (2) | 0 (0) | NR | 0 (0) | 0 (0) | Low |
| Lin, 2020 (8) | Case series | China | ICU and non-ICU | 92 | Adults | NR | NR | NR | NR | 6 (7) | NR | NR | Medium |
| Liu, 2020 (48) | Retrospective cohort | China | ICU and non-ICU | 12 | Children/Adults | 66 | NR | 6 (50) | NR | 0 (0) | 2 (17) | 0 (0) | Low |
| Lv, 2020 (49) | Retrospective cohort | China | ICU and non-ICU | 354 | Adults | 49 | NR | NR | 11 (3) | 1 (0.3) | 32 (9) | 6 (2) | Low |
| Ma, 2020 (50) | Retrospective cohort | China | NR | 93 | Adults | 55 | NR | NR | 44 (47) | 46 (49) | 0 (0) | 0 (0) | Low |
| Mannheim, 2020 (51) | Case series | USA | ICU and non-ICU | 64 | Children | 56 | 11 | NR | 0 (0) | 3 (5) | 1 (2) | 0 (0) | Medium |
| Mo, 2020 (52) | Case series | China | ICU and non-ICU | 155 | Adults | 55 | NR | 36 (23) | 22 (14) | 13 (8) | 2 (1) | 0 (0) | Medium |
| Nowak, 2020 (9) | Case series | USA | ICU and non-ICU | 1204 | Adults | 56 | NR | NR | NR | 36 (3) | 0 (0) | 0 (0) | Medium |
| Ozaras, 2020 (53) | Case series | Turkey | ICU and non-ICU | 1103 | Adults | 50 | NR | NR | NR | 6 (0.5) | 0 (0) | 0 (0) | Medium |
| Palmieri, 2020 (54) | Retrospective cohort | Italy | ICU and non-ICU | 3032 | Children/Adults | 67 | NR | NR | 3032 (100) | NR | NR | NR | Low |
| Peng, 2020 (55) | Retrospective cohort | China | ICU and non-ICU | 75 | Children | 58 | NR | NR | 0 (0) | 8 (11) | 31 (41) | 0 (0) | Low |
| Pongpirul, 2020 (56) | Case series | Thailand | ICU and non-ICU | 11 | Adults | 54 | NR | 0 (0) | 0 (0) | 2 (18) | 5 (45) | 0 (0) | Low |
| Richardson , 2020 (57) | Case series | USA | ICU and non-ICU | 5700 | Adults | 60 | 14.2 | 1151 (20) | 553 (10) | 39 (0.7) | 3 (0.1) | 0 (0) | Low |
| Sun, 2020 (58) | Retrospective cohort | China | ICU and non-ICU | 36 | Children | 61 | NR | NR | 1 (3) | 1 (3) | 1 (3) | 0 (0) | Medium |
| Tagarro, 2020 (59) | Retrospective cohort | Spain | ICU and non-ICU | 41 | Children | 44 | 9.7 | 4 (10) | 0 (0) | 2 (5) | 0 (0) | 0 (0) | Low |
| Wan, 2020 (60) | Case series | China | ICU and non-ICU | 135 | Adults | 53 | NR | 28 (21) | 1 (0.7) | NR | NR | NR | Medium |
| Wang Y, 2020 (61) | Case series | China | ICU and non-ICU | 55 | Adults | 40 | 0 | 0 (0) | 0 (0) | 1 (2) | 1 (2) | 1 (3) | Low |
| Wang L, 2020 (62) | Case series | China | ICU and non-ICU | 339 | Adults | 49 | NR | NR | 65 (19) | 0 (0) | 1 (0.3) | 1 (0.3) | Low |
| Wang R, 2020 (63) | Case series | China | ICU and non-ICU | 125 | Adults | 56.8 | 15.2 | 4 | 0 (0) | 1 (0.8) | 9 (7) | 9 (7) | Medium |
| Wang Y, 2020 (21) | Clinical trial | China | ICU and non-ICU | 237 | Adults | 56 | NR | 21 (9) | 14 (6) | NR | NR | NR | Medium |
| Wee, 2020 (64) | Prospective cohort | Singapore | ICU and non-ICU | 3807 | Adults | NR | NR | NR | 1 (0.02) | 3 (0.08) | NR | NR | Medium |
| Wu C, 2020 (65) | Retrospective cohort | China | ICU and non-ICU | 201 | Adults | 63.7 | 26.4 | 67 (33) | 44 (22) | 1 (0.5) | 0 (0) | 0 (0) | Low |
| Xia, 2020 (66) | Case series | China | ICU and non-ICU | 20 | pediatric | 65 | NR | 0 (0) | 0 (0) | 4 (0.2) | 1 (5) | 1 (5) | Medium |
| Yang X, 2020 (67) | Case series | China | ICU | 710 | Adults | 67 | 100 | 37 (5) | 32 (4) | 0 (0) | 4 (0.6) | 4 (0.6) | Low |
| Yi, 2020 (68) | Case series | USA | ICU and non-ICU | 132 | Adult | 62 | 50 | 5 (4) | 1 (0.8) | NR | NR | NR | Medium |
| Zhang J, 2020 (69) | Case series | China | ICU and non-ICU | 140 | Adults | 50.7 | NR | NR | NR | 2 (1) | 1 (0.7) | 1 (0.7) | Medium |
| Zhang G, 2020 (23) | Case series | China | ICU and non-ICU | 221 | Adults | 48.9 | 80 | 26 (12) | 5 (2) | 2 (0.9) | 6 (3) | 6 (3) | Medium |
| Zhao, 2020 (70) | Case series | China | ICU and non-ICU | 34 | Adults | 57.9 | 0 | 0 (0) | 0 (0) | 1 (3) | 1 (3) | 0 (0) | Medium |
| Zheng, 2020 (71) | Case series | China | ICU and non-ICU | 1001 | Adult and pediatric | NR | NR | NR | NR | 2 (0.2) | NR | NR | Low |
| Zhou, 2020 (72) | Retrospective cohort | China | ICU and non-ICU | 191 | Adult | 62 | 26 | 32 (17) | 54 (28) | NR | NR | NR | Low |
| Zhu, 2020 (27) | Retrospective cohort | China | ICU and non-ICU | 257 | Adult and pediatric | 53.7 | 1.16 | 0 (0) | 0 (0) | 9 (3) | 11 (4) | 11 (4) | Low |

<sup>a</sup>ICU: intensive care unit.
<sup>b</sup>NR: Not reported.

The pooled prevalence of co-infection was 12% (95% confidence interval (CI): 6%-18%, n=29, I^2^=98%). The highest prevalence of co-infection was observed among non-ICU patients as 28% (95% CI: 6%-56%), while it was 10% (95% CI: 5%-16%) among combined ICU and non-ICU patients, and 3% (95% CI: 0%-1%) among only ICU co-infected patients (Figure 2). The pooled prevalence of super-infection was 14% (95% CI: 9%-21%), with the highest prevalence of 17% (95% CI: 1%-43%) among ICU patients (Figure 3).

**Figure 2.**
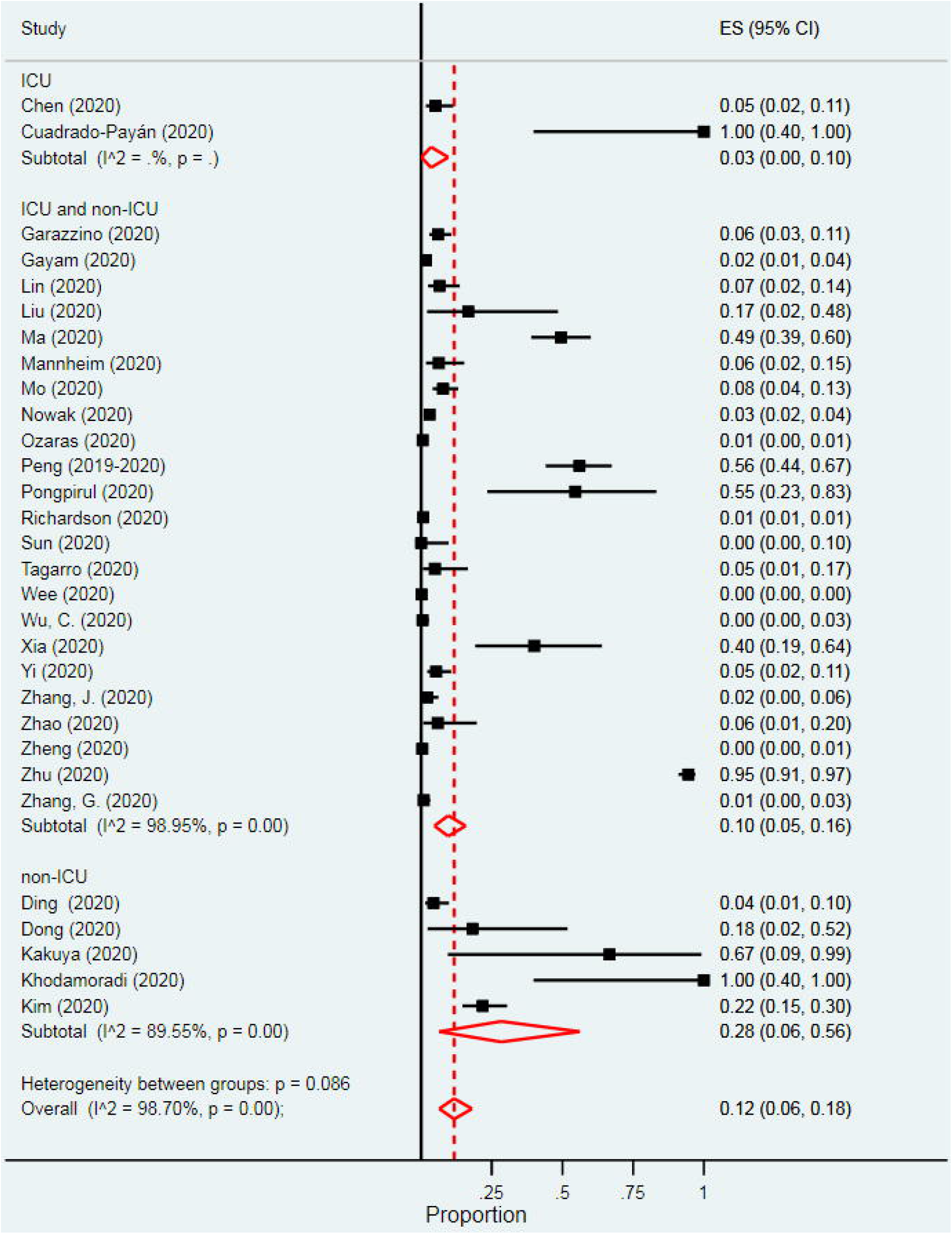
Pooled prevalence of co-infection in patients infected with SARS-CoV-2.

**Figure 3.**
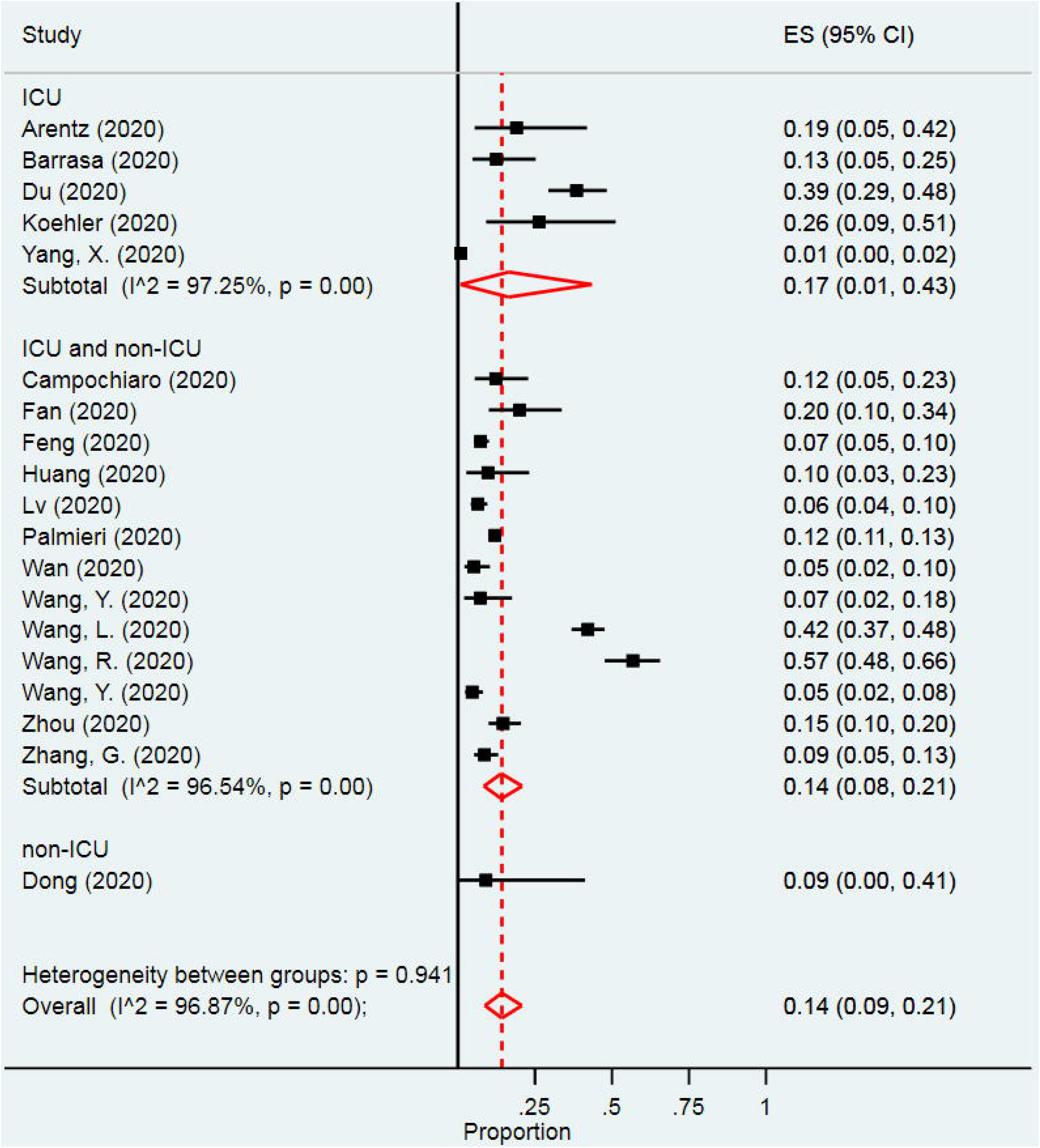
Pooled prevalence of super-infection in patients infected with SARS-CoV-2.

Pooled prevalence of pathogen type stratified by co- or super-infection was: viral co-infections, 4% (95% CI: 2%-7%); viral super-infections, 2% (95% CI: 0%-7%); bacterial co-infections, 4% (95% CI: 1%-8%); bacterial super-infections, 6% (95% CI: 2%-11%); fungal co-infections, 4% (95% CI: 1%-8%); and fungal super-infections, 4% (95% CI: 0%-11%) (S1 Fig, S2 Fig, S3 Fig).

Twenty-nine studies reported data on specific organisms associated with co-infection or super-infection in COVID-19 patients (Table 2). Among those with co-infections, the three most frequently identified bacteria were *Streptococcus pneumoniae* (17.9%), *Klebsiella pneumoniae* (16.7%) and *Haemophilus influenza* (12.4%). The three most frequently identified viruses among co-infected patients were *influenza* type A (8.1%), *Rhinovirus* (6.3%) and non-SARS-CoV-2 coronaviruses (3.7%). For fungi, only *Candida sp*. (0.7%) and *Mucor sp*. (0.7%) were identified among those co-infected.

**Table 2.** All identified organisms as a proportion of total number of organisms per pathogen type by co-infection and super-infection.

| **Pathogen type** | **Co-infection**<br>**(N= 860)** | **Super-**<br>**infection**<br>**(N=50)** |
| --- | --- | --- |
| **Bacteria, No. (%)** |  |  |
| *Staphylococcus aureus* | 22 (2.6) | 5 (10) |
| *Haemophilus influenza* | 107 (12.4) | 1 (2) |
| *Mycoplasma pneumoniae* | 47 (5.5) | 3 (6) |
| *Acinetobacter spp* | 68 (7.9) | 11 (22) |
| *Escherichia coli* | 25 (2.9) | 9 (18) |
| *Stenotrophomonas maltophilia* | 0 (0) | 2 (4) |
| *Klebsiella pneumoniae* | 144 (16.7) | 2 (4) |
| *Streptococcus pneumoniae* | 154 (17.9) | 0 (0) |
| *Chlamydia pneumoniae* | 6 (0.7) | 0 (0) |
| *Bordetella* | 3 (0.3) | 0 (0) |
| *Moraxella catarrhalis* | 11 (1.3) | 0 (0) |
| *Pseudomonas* | 12 (1.4) | 8 (16) |
| *Enterococcus faecium* | 3 (0.3) | 0 (0) |
| **Viruses, No. (%)** |  |  |
| Non-SARS-CoV-2<sup>a</sup> coronavirus strains | 32 (3.7) | 0 (0) |
| Human influenza A | 70 (8.1) | 0 (0) |
| Human influenza B | 27 (3.1) | 0 (0) |
| Respiratory syncytial virus | 25 (2.9) | 0 (0) |
| Parainfluenza | 5 (0.6) | 0 (0) |
| Human metapneumovirus | 12 (1.4) | 2 (4) |
| Rhinovirus | 54 (6.3) | 0 (0) |
| Adenovirus | 21 (2.4) | 0 (0) |
| **Fungus, No. (%)** |  |  |
| *Mucor* | 6 (0.7) | 0 (0) |
| *Candida spp* | 6 (0.7) | 7 (14) |

<sup>a</sup>SARS-CoV-2: severe acute respiratory syndrome coronavirus 2

Among those with super-infections, the three most frequently identified bacteria were *Acinetobacter spp*. (22%), *Escherichia coli* (18%) and *Pseudomonas* (16%). For viruses, only Human *metapneumovirus* (4%) was identified among those super-infections and for fungi only *Candida sp*. (14%) was identified.

Two studies reported both super-infections and co-infections with SARS-CoV-2 for the same population.(22, 23) These warrant further mention. In Zhang G et. al., super-infections were due to bacterial respiratory infections and 25% (14/55) of the patients had severe disease. Stratification by disease severity for mortality was not reported for these super-infected patients. Co-infections were due to non-specified viruses and of those patients, 29% (16/55) had severe COVID-19. In Dong et. al., one (1/11) patient had both a bacterial super-infection and viral co-infection with severe COVID-19.

The overall prevalence of comorbidities was 45% (95% CI: 33%-56%). Among those with co-infections the prevalence of comorbidities was 38% (95% CI: 24%-53%), while it was 54% (95% CI: 35%-72%) among those who were super-infected.

COVID-19 patients with a co-infection or super-infection had higher odds of dying than those who only had COVID-19 infection, but this was not statistically significant (odds ratio [OR] 2.59, 95% CI: 0.83-8.09). Subgroup analysis of mortality showed similar results where the odds of death were higher among patients who were co-infected and those who were super-infected, but the findings were not statistically significant, OR 1.96: (95% CI: 0.62-6.2) and OR 3.2: (95% CI: 0.54-18.3), respectively. There was a higher prevalence of mechanical ventilation among patients with super-infections, 21% (95% CI: 13%-31%) compared to those with co-infections, 7% (95% CI: 2%-15%).

Twenty-five studies reported data on average LOS. The average LOS for co-infected patients was 10.2 days, standard deviation (SD) = 6.7, while the average LOS for super-infected patients was 12.5 days, SD=5.3. None of the studies included in this meta-analysis reported data on discharge disposition and readmissions. There was asymmetry of the funnel plot which suggests publication bias (S3 Fig).

### Risk of bias assessment

Forty-seven percent (23/48) of studies were rated as having low risk of bias, 50% (24/48) as having medium risk of bias and 1 study was rated as having a high risk of bias (24) (S1 Table).

## DISCUSSION

We found that 12% of SARS-CoV-2 patients were co-infected with other pathogens and the prevalence of co-infection was higher among patients who were not in the ICU (28%). We also found a higher prevalence of super-infection compared to co-infection (14%), particularly among ICU patients (17%). Further, we found that super-infected patients had a higher prevalence of mechanical ventilation, ICU admission and comorbidities. Super-infected patients had higher odds of death, although this was not statistically significant.

Two previous reviews that have focused on this question found similar prevalence of bacterial co-infection in SARS-CoV-2 infected patients of 7% - 8% and viral co-infection of 3%.(11, 12) We extended this work by distinguishing between super- and co-infection because of the different implications of co-infections vs super-infections. In particular, bacterial and other pathogens have been shown to complicate viral pneumonia and lead to poor outcomes.(25)

The three most frequently identified bacteria among co-infected patients in our study were *Streptococcus pneumoniae, Klebsiella pneumoniae* and *Haemophilus influenza. Streptococcus pneumoniae* is a frequent cause of super-infection in other respiratory infections like influenza.(26) A study by Zhu et al. showed similar results,(27) and Lansbury et al. showed that *Klebsiella pneumoniae* and *Haemophilus influenza* were some of the most frequent bacterial co-infecting pathogens identified in their review.(11) As expected, *Staphylococcus aureus* also was present in a sizeable number of cases. The most frequent bacteria identified in super-infected patients was *Acinetobacter spp*., which is a common infection especially in ventilated patients.(28)

In our study, the three most frequently identified viruses among co-infected patients were *Influenza type A, Rhinovirus* and non-SARS-CoV-2 coronaviruses. These findings are important particularly for influenza because testing constraints continue to exist, yet clinical presentation of influenza and SARS-CoV-2 is similar. There are major infection control and clinical implications of missing SARS-CoV-2 or influenza if co-infection is not considered, and diagnostic testing for both pathogens is not undertaken. The only identified virus among patients with super-infections was Human *metapneumovirus (hMPV)*. This may have clinical implications as both viruses— hMPV and SARS-CoV- 2—share poor prognosis-associated factors such as advanced age and being immunocompromised.(29, 30)

Our findings have implications for infection preventionists, clinicians and laboratory leaders. While it is not possible to predict with certainty how the upcoming respiratory virus season will unfold, it is likely that we will be dealing with a multitude of viruses circulating simultaneously. Respiratory virus diagnostic testing protocols should take into account that co-infection with SARS-CoV-2 is not infrequent and therefore viral panel testing may be advisable in patients with compatible symptoms. Treatment protocols should also include assessing for co-infections, particularly influenza, so that appropriate treatment for both SARS-CoV-2 and Influenza can be administered.

Our study has limitations. We were not able to assess important outcomes such as discharge disposition and hospital readmissions due to lack of availability of this data in the included studies. We were not able to document time to super-infection as included studies did not report this information. Studies provided the number of patients with super-infections without stating the exact time when this determination was made after SARS-COV-2 diagnosis. Most of the studies included in the meta-analysis were case series with their inherent limitations.(31) It is possible that some of the pathogens that were reported as super-infections or secondary infections were present but not tested for at admission and hence were co-infections. It was not possible to assess this from the studies. There was significant heterogeneity in the studies, as was anticipated given the variation in settings, patient populations and diagnostic testing platforms across the studies.

## CONCLUSIONS

In conclusion, our study shows that as many as 14% patients with COVID-19 have super-infections and 12% have co-infections. Poor outcomes were associated with super-infections. Our results have implications for diagnostic testing, laboratory and antibiotic stewardship, particularly with the upcoming respiratory virus season in the fall.

## Data Availability

All data used in analysis of this manuscript is freely available by contacting the corresponding author.

