## Supplementary figures and images for "Prevalence and outcomes of co-infection and super-infection with SARS-CoV-2 and other pathogens: A Systematic Review and Meta-analysis"

### Supplemental Figure 1

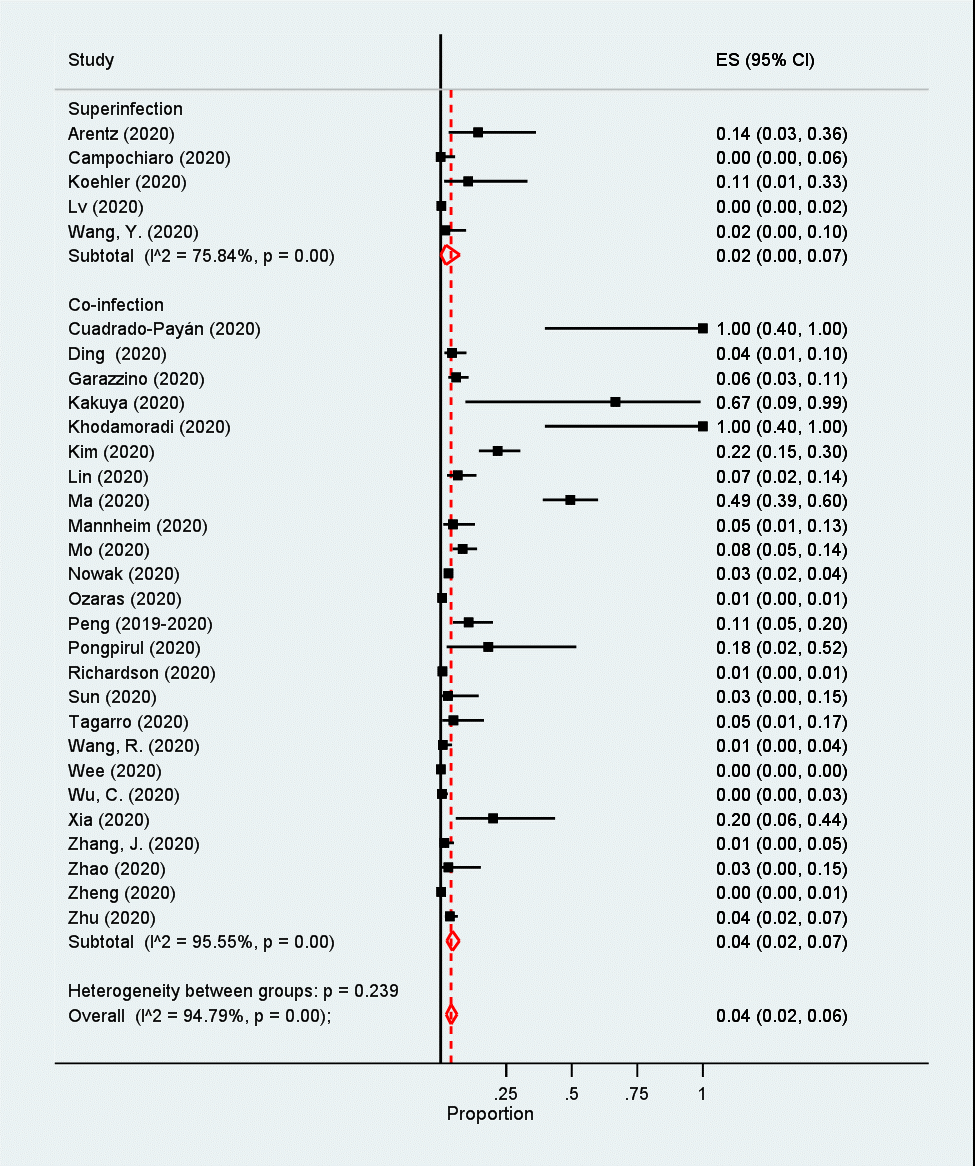

### Supplemental Figure 2

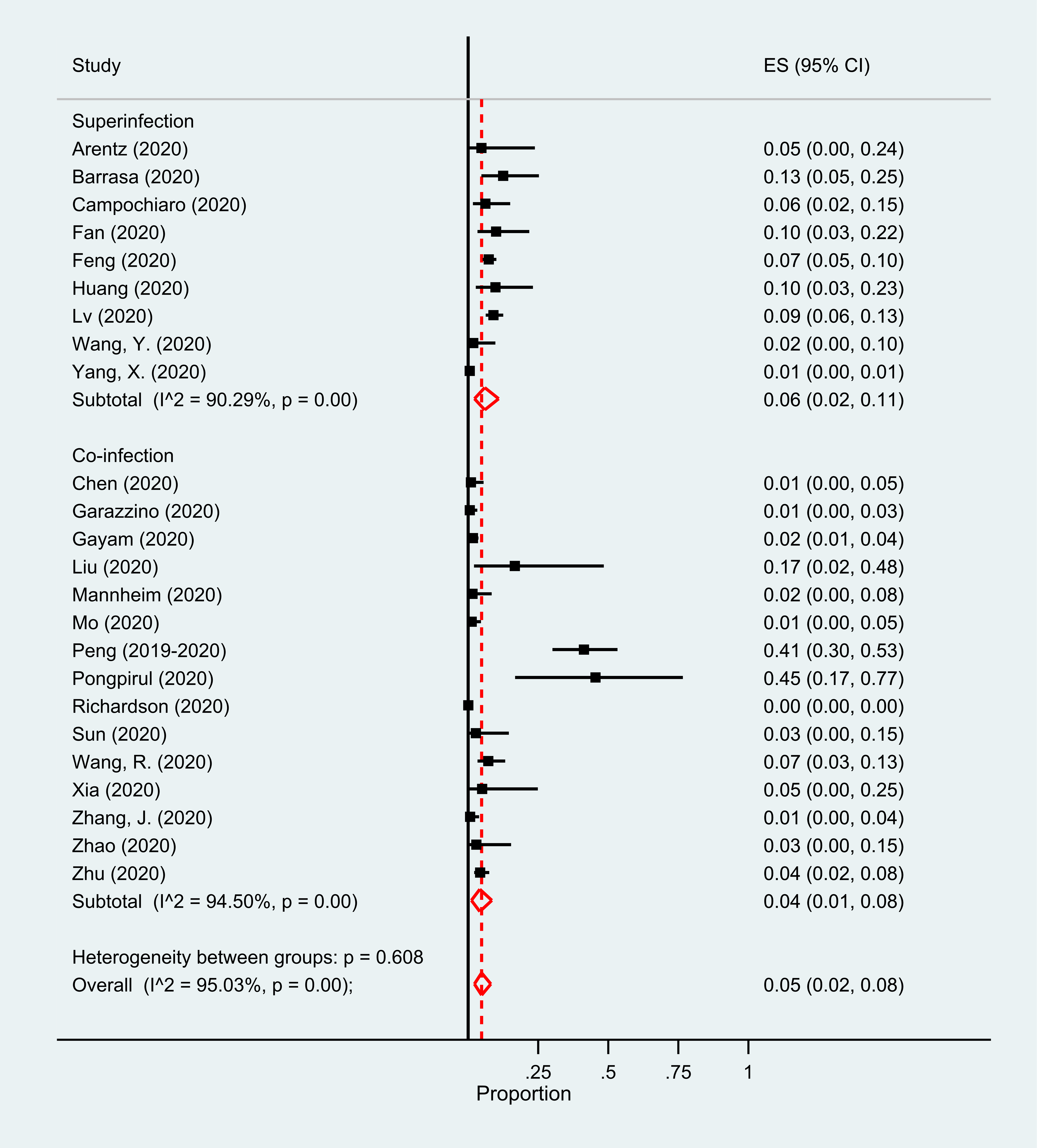

### Supplemental Figure 3

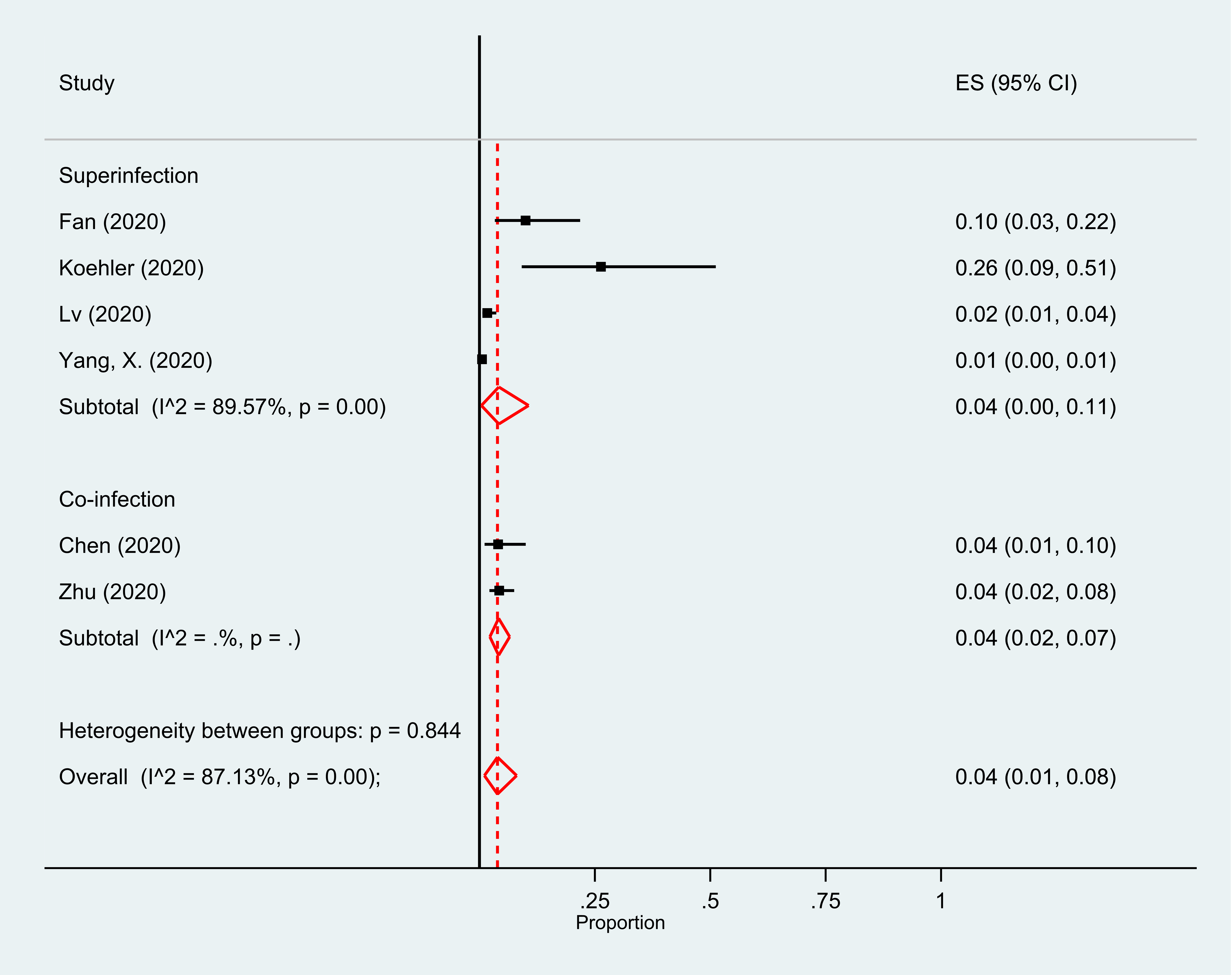
