## Supplemental Table 1 for "Prevalence and outcomes of co-infection and super-infection with SARS-CoV-2 and other pathogens: A Systematic Review and Meta-analysis"

**Supplement Table 1.** Risk of bias assessment

| Study | Year | Study design | 1 | 2 | 3 | 7 | 8 | Quality assessment score | Risk of bias |
|---|---|---|---|---|---|---|---|---|---|
| Wang, Y | 2020 | Clinical trial | 0 | 1 | 1 | 1 | 1 | 4 | Medium |
| Campochiaro | 2020 | Prospective cohort | 1 | 1 | 1 | 1 | 1 | 5 | Low |
| Wee | 2020 | Prospective cohort | 1 | 1 | 1 | 1 | 0 | 4 | Medium |
| Fan | 2020 | Retrospective cohort | 1 | 1 | 1 | 1 | 1 | 5 | Low |
| Garazzino | 2020 | Retrospective cohort | 1 | 1 | 1 | 1 | 1 | 5 | Low |
| Kim | 2020 | Retrospective cohort | 1 | 1 | 1 | 1 | 1 | 5 | Low |
| Lian | 2020 | Retrospective cohort | 1 | 1 | 1 | 1 | 1 | 5 | Low |
| Liu | 2020 | Retrospective cohort | 1 | 1 | 1 | 1 | 1 | 5 | Low |
| Lv | 2020 | Retrospective cohort | 1 | 1 | 1 | 1 | 1 | 5 | Low |
| Ma | 2020 | Retrospective cohort | 1 | 1 | 1 | 1 | 1 | 5 | Low |
| Palmieri | 2020 | Retrospective cohort | 1 | 1 | 1 | 1 | 1 | 5 | Low |
| Peng | 2020 | Retrospective cohort | 1 | 1 | 1 | 1 | 1 | 5 | Low |
| Sun | 2020 | Retrospective cohort | 0 | 1 | 1 | 1 | 1 | 4 | Medium |
| Tagarro | 2020 | Retrospective cohort | 1 | 1 | 1 | 1 | 1 | 5 | Low |
| Wu, C. | 2020 | Retrospective cohort | 1 | 1 | 1 | 1 | 1 | 5 | Low |
| Zhou | 2020 | Retrospective cohort | 1 | 1 | 1 | 1 | 1 | 5 | Low |
| Zhu | 2020 | Retrospective cohort | 1 | 1 | 1 | 1 | 1 | 5 | Low |
| Arentz | 2020 | Case series | 1 | 1 | 1 | 0 | 1 | 4 | Medium |
| Barrasa | 2020 | Case series | 1 | 1 | 1 | 1 | 1 | 5 | Low |
| Chen | 2020 | Case series | 1 | 1 | 1 | 0 | 1 | 4 | Medium |
| Cuadrado-Payán | 2020 | Case series | Unable to determine | 1 | 0 | 0 | 1 | 2 | High |
| Ding | 2020 | Case series | 1 | 1 | 1 | Unable to | 1 | 4 | Medium |

| | | | | | | | | | |
|---|---|---|---|---|---|---|---|---|---|
| | | | | | | deter mine | | | |
| Dong | 2020 | Case series | 0 | 1 | 0 | 1 | 1 | 3 | Medium |
| Du | 2020 | Case series | 1 | 1 | 1 | 1 | 1 | 5 | Low |
| Feng | 2020 | Case series | 1 | 1 | 1 | 0 | 1 | 4 | Medium |
| Gayam | 2020 | Case series | 1 | 1 | 1 | 0 | 1 | 4 | Medium |
| Huang | 2020 | Case series | 1 | 1 | 1 | 0 | 1 | 4 | Medium |
| Kakuya | 2020 | Case series | 1 | 1 | 1 | 1 | 1 | 5 | Low |
| Khodamoradi | 2020 | Case series | 1 | 1 | 1 | 0 | 1 | 4 | Medium |
| Koehler | 2020 | Case series | 1 | 1 | 0 | 0 | 1 | 3 | Medium |
| Lin | 2020 | Case series | 0 | 1 | 1 | 0 | 1 | 3 | Medium |
| Mannheim | 2020 | Case series | 1 | 1 | 1 | 0 | 1 | 4 | Medium |
| Mo | 2020 | Case series | 1 | 1 | 1 | 0 | 1 | 4 | Medium |
| Nowak | 2020 | Case series | 1 | 1 | 1 | 0 | 1 | 4 | Medium |
| Ozaras | 2020 | Case series | 1 | 0 | 1 | 0 | 1 | 3 | Medium |
| Pongpirul | 2020 | Case series | 1 | 1 | 1 | 1 | 1 | 5 | Low |
| Richardson | 2020 | Case series | 1 | 1 | 1 | 1 | 1 | 5 | Low |
| Wan | 2020 | Case series | 1 | 1 | 1 | 0 | 1 | 4 | Medium |
| Wang, Y. | 2020 | Case series | 1 | 1 | 1 | 1 | 1 | 5 | Low |
| Wang, L. | 2020 | Case series | 1 | 1 | 1 | 1 | 1 | 5 | Low |
| Wang, R. | 2020 | Case series | 0 | 1 | 1 | 0 | 1 | 3 | Medium |
| Xia | 2020 | Case series | 0 | 1 | 1 | 1 | 0 | 3 | Medium |
| Yang, X. | 2020 | Case series | 1 | 1 | 1 | 1 | 1 | 5 | Low |
| Yi | 2020 | Case series | 0 | 1 | 1 | 1 | 1 | 4 | Medium |
| Zhang, J. | 2020 | Case series | 0 | 1 | 1 | 0 | 1 | 3 | Medium |
| Zhang, G. | 2020 | Case series | 0 | 1 | 1 | 0 | 1 | 3 | Medium |
| Zhao | 2020 | Case series | 0 | 1 | 1 | 0 | 1 | 3 | Medium |
| Zheng | 2020 | Case series | 1 | 1 | 1 | 1 | 1 | 5 | Low |
