## Supplemental File 1 for "Prevalence and outcomes of co-infection and super-infection with SARS-CoV-2 and other pathogens: A Systematic Review and Meta-analysis"

### Citation

Jackson Musuuza, Nasia Safdar, Lauren Watson, Vishala Parmasad, Nathan Putman-Buehler, Leslie Christensen. The prevalence and outcomes of co-infection with COVID-19 and other pathogens: a rapid systematic review and meta-analysis. PROSPERO 2020 CRD42020189763 Available from: https://www.crd.york.ac.uk/prospero/display_record.php?ID=CRD42020189763

### Review question

What is the prevalence of co-infections, and what are the associated outcomes, among patients with COVID-19?

### Searches

We will perform searches using the following online databases: the Cochrane Central Register of Controlled Trials (CENTRAL), PubMed, CINAHL, Scopus (including EMBASE), and Web of Science.

We will develop word combinations for the searches from a list of words organized into three categories: COVID-19, co-infections, outcomes.

We will exclude abstracts, reviews, case reports, editorials, qualitative studies and articles in languages other than English.

### Types of study to be included

Randomized controlled studies, observational studies, and quasi-experimental designs.

### Condition or domain being studied

COVID-19.

### Participants/population

All COVID-19 positive patients with co-infections.

### Intervention(s), exposure(s)

Not applicable (this review is looking at the prevalence and outcomes of co-infection with COVID-19 and other pathogens).

### Comparator(s)/control

Not applicable.

### Main outcome(s)

The proportion of patients with co-infections.

#### * Measures of effect

Proportions/odds ratios.

### Additional outcome(s)

Outcomes associated with co-infections: ICU admission, death, etc.

#### * Measures of effect

Proportions/relative risks.

### Data extraction (selection and coding)

Two reviewers (JSM and LW) will independently extract data from individual studies using a predefined template. We will collect data on: study design/methodology, location and setting (intensive care or ICU, in patient non-ICU, or outpatient, where applicable), study population, proportion of patients with co-infections, implicated pathogens, method of detection of co-infections (laboratory verified or clinical features only), type of infection— bacterial, viral or fungal, outcomes of co-infected patients— death, ICU admission, mechanical ventilation, discharge disposition, length of hospital stay, or mild illness. Data will be extracted in duplicate using Covidence (https://www.covidence.org/). Discrepancies will be discussed between the two abstractors and a third reviewer (NS) will arbitrate if necessary.

### Risk of bias (quality) assessment

We will use the Downs and Black tool to assess for bias in the included studies.

### Strategy for data synthesis

We will conduct a meta-analysis to estimate the proportion of patients co-infected with either bacterial, viral or fungal infection. We will calculate the proportion of patients co-infected with specific pathogens. Given the novelty of the main disease studied (COVID-19) and the differences in location of studies, we anticipate a high-level of heterogeneity. Regardless, we will proceed to conduct the meta-analysis given the importance of the question. We will use the “meta” command in STATA to estimate the pooled proportion of co-infected patients using a random effects model (DerSimonian Laird). In order to correctly estimate extreme proportions (i.e., those close to 0% or 100%), we will stabilize variance using an appropriate method such as the Freeman-Tukey arcsine transformation. We will assess heterogeneity using the $I^2$ statistic.

### Analysis of subgroups or subsets

Subgroup analyses will be conducted for:

ICU patients only vs. mixed hospitalized populations;
Age groups;

Location/country.

### Contact details for further information

Jackson Musuuza


### Organisational affiliation of the review

University of Wisconsin, Madison

### Review team members and their organisational affiliations

Dr Jackson Musuuza. University of Wisconsin, Madison
Dr Nasia Safdar. William S. Middleton Memorial Veteran’s Hospital & Department of Medicine, University of Wisconsin School of Medicine and Public Health
Mrs Lauren Watson. Department of Medicine, University of Wisconsin School of Medicine and Public Health
Dr Vishala Parmasad. Department of Medicine, University of Wisconsin School of Medicine and Public Health
Mr Nathan Putman-Buehler. Department of Medicine, University of Wisconsin School of Medicine and Public Health
Leslie Christensen. Ebling Library for the Health Sciences

### Type and method of review

Epidemiologic, Meta-analysis, Narrative synthesis, Prognostic, Systematic review, Other

### Anticipated or actual start date

02 June 2020

### Anticipated completion date

31 July 2020

### Funding sources/sponsors

None

### Conflicts of interest

### Language
English

### Country
United States of America

### Stage of review
Review Ongoing

### Subject index terms status
Subject indexing assigned by CRD

### Subject index terms
Bacterial Infections; Coinfection; Coronavirus; Coronavirus Infections; COVID-19; Disease Progression; Humans; Mortality; Prevalence; Prognosis; Risk; Risk Factors; Virus Diseases

### Date of registration in PROSPERO
03 June 2020

### Date of first submission
01 June 2020

### Stage of review at time of this submission

| Stage | Started | Completed |
|---|---|---|
| Preliminary searches | Yes | No |
| Piloting of the study selection process | No | No |
| Formal screening of search results against eligibility criteria | No | No |
| Data extraction | No | No |
| Risk of bias (quality) assessment | No | No |
| Data analysis | No | No |

### Revision note
After discussions with the study team, we decided to include letters and correspondences as long as they have data on COVID-19 and co-infections.

*The record owner confirms that the information they have supplied for this submission is accurate and complete and they understand that deliberate provision of inaccurate information or omission of data may be construed as scientific misconduct.*

*The record owner confirms that they will update the status of the review when it is completed and will add publication details in due course.*

### Versions
03 June 2020

PROSPERO
This information has been provided by the named contact for this review. CRD has accepted this information in good faith and registered the review in PROSPERO. The registrant confirms that the information supplied for this submission is accurate and complete. CRD bears no responsibility or liability for the content of this registration record, any associated files or external websites.
