## Supplemental File 2 for "Prevalence and outcomes of co-infection and super-infection with SARS-CoV-2 and other pathogens: A Systematic Review and Meta-analysis"

**Supplementary material: Search Strategies.** COVID-19 and co-infections. Final Search Strategies

1820 total citations (English Language) retrieved from databases | 19 articles from hand search | 1310 records (after 529 duplicates removed)

**Pubmed**

**Search Date: 6/11/2020**

**563 Results | English (526 results)**

((“Coronavirus”[mesh] OR “Coronavirus Infections”[mesh] OR coronavir*[tw] OR “corona vir*”[tw] OR OC43[all] OR NL63[all] OR 229E[all] OR HKU1[all] OR HCoV*[tw] OR nCoV*[tw] OR covid*[tw] OR SARS*[tw] OR “Severe Acute Respiratory Syndrome Coronavir*”[tw] OR ((pneumonia[mesh] OR pneumonia*[tw] OR “Influenza, Human”[mesh] OR influenza*[tw] OR flu[tw]) AND (wuhan[tiab] OR hubei[tiab])) AND 2019/10[pdat]:2030[pdat])) OR ("COVID-19"[Supplementary Concept] OR "Severe Acute Respiratory Syndrome Coronavirus 2"[Supplementary Concept] OR “2019 nCoV”[tw] OR nCoV19[tw] OR “nCoV 19”[tw] OR “SARS CoV2"[tw] OR “SARS CoV 2”[tw] OR SARSCoV2[tw]OR “SARSCoV 2”[tw] OR “SARS coronavirus2”[tw] OR “SARS coronavirus 2”[tw] OR “SARS like coronavir*”[tw] OR “severe acute respiratory syndrome coronavirus 2”[tw] OR “coronavirus 19”[tw] OR covid19[tw] OR “covid 19”[tw] OR covid2019[tw] OR “covid 2019”[tw] OR nCoV2019[tw] OR “nCoV 2019”[tw] OR (“2019 novel”[tw] AND (coronavir* OR “corona vir*”[tw] OR CoV[tw] OR nCOV[tw] OR covid[tw]))) AND (“Coinfection”[mesh] OR “Superinfection”[mesh] OR superinfect*[tw] OR “super infect*”[tw] OR coinfect*[tw] OR “co infect*”[tw] OR ((bacteria*[tw] OR fungus[tw] OR fungal[tw] OR mycoses[tw] OR secondary[tw] OR concomitant[tw] OR mixed[tw]) AND infect*[tw]) OR “Pneumovirinae”[mesh] OR “Pneumovirus Infections”[mesh] OR pneumovir*[tw] OR "Respiratory Syncytial Viruses"[Mesh] OR “respiratory syncytial virus*”[tw] OR RSV[tw] OR “Metapneumovirus”[mesh] OR metapneumovir*[tw] OR “meta pneumovir*”[tw] OR HMPV[tw] OR “Influenza, Human”[mesh] OR “Influenzavirus A”[mesh] OR influenza*[tw] OR “respiratory virus*”[tw] OR “Bacterial Infections and Mycoses”[mesh] OR (“upper respiratory”[tw] AND (infection*[tw] OR virus*[tw]))) AND (“Respiration, Artificial”[Mesh] OR (artificial[tw] AND respiration*[tw]) OR oxygen[tw] OR “Oxygen Inhalation Therapy”[mesh] OR “Intensive Care Units”[mesh] OR “intensive care”[tw] OR ICU OR ICUs[tw] OR “Critical Care”[mesh] OR “critical care”[tw] OR critical*[tiab] OR ventilat*[tw] OR intubat*[tw] OR outcome*[tw] OR “Nursing Homes”[mesh] OR “Subacute Care”[mesh] OR “nursing home*”[tw] OR “skilled nursing”[tw] OR “intermediate care”[tw] OR “Patient Discharge”[mesh] OR “Mortality”[mesh] OR “Mortality”[subheading] OR mortalit*[tw] OR “Morbidity”[mesh] OR death*[tw] OR morbidit*[tw] OR (patient*[tw] AND discharg*[tw])) AND English[filter]

-----

**Scopus**

**Search Date: 6/11/2020**

**651 results | English Language (619 results)**

((TITLE-ABS-KEY(coronavir* OR "corona vir*" OR OC43 OR NL63 OR 229E OR HKU1 OR HCoV* OR ncov* OR covid* OR sars* OR "Severe Acute Respiratory Syndrome")) OR (TITLE-ABS-KEY((pneumonia* OR influenza* OR flu) AND (wuhan OR hubei))) AND (( PUBDATETXT (“october 2019” OR "november 2019" OR "december 2019" ) ) OR (PUBYEAR > 2019))) OR (TITLE-ABS-KEY("Severe Acute Respiratory Syndrome Coronavirus 2" OR "2019 ncov" OR ncov19 OR "ncov 19" OR "2019 novel CoV" OR "sars cov2" OR "SARS CoV 2" OR SARSCoV2 OR "SARSCoV 2" OR "SARS coronavirus2" OR "SARS coronavirus 2" OR "SARS like coronavir*" OR "coronavirus 19" OR covid19 OR "covid 19" OR covid2019 OR "covid 2019" OR nCoV2019 OR "nCoV 2019")) AND ( TITLE-ABS-KEY ( coinfect* OR "co infect*" OR superinfect* OR “super infect*”) ) OR ( TITLE-ABS-KEY ( ( bacteria* OR fungus OR fungal OR mycoses OR secondary OR concomitant OR mixed ) AND infect* ) ) OR ( TITLE-ABS-KEY ( pneumovir* OR "respiratory syncytial virus*" OR rsv OR metapneumovir* OR “meta pneumovir*” OR hmpv OR influenza* OR (respiratory W/2 virus*) )) OR ( TITLE-ABS-KEY ( ( upper W/3 respiratory ) AND ( infection* OR virus* ) ) ) AND TITLE-ABS-KEY((artificial AND respiration*) OR oxygen OR "intensive care" OR ICU OR ICUs OR "critical care" OR ventilat* OR intubat* OR outcome* OR "nursing home*" OR "skilled nursing" OR “subacute care” OR "intermediate care" OR mortalit* OR death* OR morbidit* OR (patient* AND discharg*)) OR TITLE-ABS(critical*) AND ( LIMIT-TO ( LANGUAGE , "English" ) )

-----

**Web of Science**

**Search Date: 6/11/2020**

**314 Results | English Language (303 results)**

(((TS=(coronavir* OR "corona vir*" OR OC43 OR NL63 OR 229E OR HKU1 OR HCoV* OR ncov* OR covid* OR sars* OR "Severe Acute Respiratory Syndrome") OR TS=((pneumonia* OR influenza* OR flu) AND (wuhan OR hubei) )) AND PY=(2019 OR 2020)) OR (TS=("Severe Acute Respiratory Syndrome Coronavirus 2" OR "2019 ncov" OR ncov19 OR "ncov 19" OR "2019 novel CoV" OR "sars cov2" OR "SARS CoV 2" OR SARSCoV2 OR "SARSCoV 2" OR "SARS coronavirus2" OR "SARS coronavirus 2" OR "SARS like coronavir*" OR "coronavirus 19" OR covid19 OR "covid 19" OR covid2019 OR "covid 2019" OR nCoV2019 OR "nCoV 2019") )) AND ((TS=(coinfect* OR "co infect*" OR superinfect* OR “super infect*”) ) OR (TS=(bacteria* OR fungus OR fungal OR mycoses OR secondary OR concomitant OR mixed ) AND TS=(infect*)) OR (TS=(pneumovir* OR "respiratory syncytial virus*" OR rsv OR metapneumovir* OR “meta pneumovir*” OR hmpv OR influenza* OR (respiratory NEAR/2 virus*) )) OR (TS=("upper respiratory" AND (infection* OR virus*) ))) AND ((TS=(artificial AND respiration*) ) OR (TS=( oxygen OR "intensive care" OR ICU OR ICUs OR "critical care" OR ventilat* OR intubat* OR outcome* OR "nursing home*" OR "skilled nursing" OR “subacute care” OR "intermediate care" OR mortalit* OR death* OR morbidit*) ) OR (TS=(patient AND discharg*) ) OR (TI=(critical*) ) OR (AB=(critical*))) AND (LA=(English))

-----

**CINAHL Plus**

**Search Date: 6/11/2020**

**255 Results | English (253 Results)**

**Search History**

| | |
|---|---|
| S30 | S25 AND S26 AND S28 Limiters - English |
| S29 | S25 AND S26 AND S28 |
| S28 | S15 OR S16 OR S17 OR S18 OR S19 OR S20 OR S21 OR S22 OR S23 OR S24 OR S27 |
| S27 | TX ( (artificial AND respiration*) OR oxygen OR "intensive care" OR ICU OR ICUs OR "critical care" OR ventilat* OR intubat* OR outcome* OR "nursing home*" OR "skilled nursing" OR “subacute care” OR "intermediate care" OR mortalit* OR death* OR morbidit* OR (patient* AND discharg*) ) OR TI critical* OR AB critical* |
| S26 | S8 OR S9 OR S10 OR S11 OR S12 OR S13 OR S14 |
| S25 | S6 OR S7 |
| S24 | (MW "mo") |
| S23 | (MH "Morbidity+") |

| | |
|---|---|
| S22 | (MH "Mortality+") |
| S21 | (MH "Patient Discharge+") |
| S20 | (MH "Subacute Care") |
| S19 | (MH "Nursing Homes+") |
| S18 | (MH "Critical Care+") |
| S17 | (MH "Intensive Care Units+") |
| S16 | (MH "Oxygen Therapy+") |
| S15 | (MH "Respiration, Artificial+") |
| S14 | TX (upper N3 respiratory) AND (infection* OR virus*) |
| S13 | TX (pneumovir* OR "respiratory syncytial virus*" OR RSV OR metapneumovir* OR "meta pneumovir*" OR HMPV OR influenza* OR "respiratory virus*") |
| S12 | (MH "Bacterial Infections+") OR (MH "Mycoses+") |
| S11 | (MH "Influenza, Human+") OR (MH "Influenzavirus A+") |
| S10 | (MH "Respiratory Syncytial Viruses") OR (MH "Respiratory Syncytial Virus Infections") |

| | |
|---|---|
| S9 | TX (bacteria* OR fungus OR fungal OR mycoses OR secondary OR concomitant OR mixed ) AND infect* |
| S8 | (MH "Coinfection") OR (MH "superinfection") OR (TX coinfect* OR "co infect* OR superinfect* OR "super infect*") |
| S7 | TX "Severe Acute Respiratory Syndrome Coronavirus 2" OR "2019 ncov" OR ncov19 OR "ncov 19" OR "2019 novel CoV" OR "sars cov2" OR "SARS CoV 2" OR SARSCoV2 OR "SARSCoV 2" OR "SARS coronavirus2" OR "SARS coronavirus 2" OR "SARS like coronavir*" OR "coronavirus 19" OR covid19 OR "covid 19" OR covid2019 OR "covid 2019" OR nCoV2019 OR "nCoV 2019" |
| S6 | S1 OR S2 OR S3 OR S4 Limiters - Published Date: 20191001- |
| S5 | S1 OR S2 OR S3 OR S4 |
| S4 | (MH "Pneumonia+" OR MH "Influenza, Human+" OR TX ( pneumonia* OR influenza* OR flu )) AND (TI ( wuhan OR hubei ) OR AB ( wuhan OR hubei )) |

| | |
|---|---|
| S3 | TX ( coronavir* OR "corona vir*" OR HCoV* OR ncov* OR covid* OR sars* OR "Severe Acute Respiratory Syndrome" ) OR ( OC43 OR NL63 OR 229E OR HKU1 ) |
| S2 | (MH "Coronavirus Infections+") |
| S1 | (MH "Coronavirus+") |

**Cochrane Central Register of Controlled Trials (CENTRAL Issue 6 of 12, June 2020)**

**Search Date: 6/11/2020**

**Search Name: Musuuza RR covid-19 and co-infection**

**118 Results**

ID Search

#1 MeSH descriptor: [Coronavirus] explode all trees

#2 MeSH descriptor: [Coronavirus Infections] explode all trees

#3 ((coronavir* OR "corona vir*" OR HCoV* OR ncov* OR covid* OR sars* OR "Severe Acute Respiratory Syndrome Coronavir*")):ti,ab,kw

#4 ((OC43 OR NL63 OR 229E OR HKU1))

#5 #1 OR #2 OR #3 OR #4

#6 MeSH descriptor: [Pneumonia] explode all trees

#7 MeSH descriptor: [Influenza, Human] explode all trees

#8 (pneumonia* OR influenza* OR flu):ti,ab,kw

#9 #6 OR #7 OR #8

#10 (wuhan OR hubei):ti

#11 (wuhan OR hubei):ab

#12 #10 OR #11

#13 #9 AND #12

#14 #5 OR #13 with Cochrane Library publication date Between Oct 2019 and Dec 2020

#15 ("Severe Acute Respiratory Syndrome Coronavirus 2" OR "2019 ncov" OR ncov19 OR "ncov 19" OR "2019 novel CoV" OR "sars cov2" OR "SARS CoV 2" OR SARSCoV2 OR "SARSCoV 2" OR "SARS coronavirus2" OR "SARS coronavirus 2" OR "SARS like coronavir*" OR "coronavirus 19" OR covid19 OR "covid 19" OR covid2019 OR "covid 2019" OR nCoV2019 OR "nCoV 2019"):ti,ab,kw

#16 #14 OR #15

#17 MeSH descriptor: [Coinfection] explode all trees

#18 (coinfect* OR "co infect*"):ti,ab,kw

#19 MeSH descriptor: [Superinfection] explode all trees

#20 (superinfect* OR "super infect*"):ti,ab,kw

#21 ((bacteria* OR fungus OR fungal OR mycoses OR secondary OR concomitant OR mixed) AND infect*):ti,ab,kw

#22 MeSH descriptor: [Pneumovirinae] explode all trees

#23 MeSH descriptor: [Respiratory Syncytial Viruses] explode all trees

#24 MeSH descriptor: [Pneumovirus Infections] explode all trees

#25 MeSH descriptor: [Metapneumovirus] explode all trees

#26 MeSH descriptor: [Influenza, Human] explode all trees

#27 MeSH descriptor: [Influenzavirus A] explode all trees

#28 MeSH descriptor: [Bacterial Infections and Mycoses] explode all trees

#29 (pneumovir* OR "respiratory syncytial virus*" OR rsv OR metapneumovir* OR “meta pneumovir*” OR hmpv OR influenza* OR (respiratory NEAR/2 virus*)):ti,ab,kw

#30 ((upper NEAR/3 respiratory) AND (infection* OR virus*)):ti,ab,kw

#31 #17 OR #18 OR #19 OR #20 OR #21 OR #22 OR #23 OR #24 OR #25 OR #26 OR #27 OR #28 OR #29 OR #30

#32 MeSH descriptor: [Respiration, Artificial] explode all trees

#33 (artificial AND respiration*):ti,ab,kw

#34 MeSH descriptor: [Oxygen Inhalation Therapy] explode all trees

#35 MeSH descriptor: [Intensive Care Units] explode all trees

#36 MeSH descriptor: [Critical Care] explode all trees

#37 MeSH descriptor: [Nursing Homes] explode all trees

#38 MeSH descriptor: [Patient Discharge] explode all trees

#39 MeSH descriptor: [Subacute Care] explode all trees

#40 MeSH descriptor: [Mortality] explode all trees

#41 MeSH descriptor: [Morbidity] explode all trees

#42 MeSH descriptor: [] explode all trees and with qualifier(s): [mortality - MO]

#43 (oxygen OR "intensive care" OR ICU OR ICUs OR "critical care" OR ventilat* OR intubat* OR outcome* OR "nursing home*" OR "subacute care" OR "skilled nursing" OR "intermediate care" OR mortalit* OR death* OR morbidit* OR (patient* AND discharg*)):ti,ab,kw

#44 (critical*):ti

#45 (critical*):ab

#46 #32 OR #33 OR #34 OR #35 OR #36 OR #37 OR #38 OR #39 OR #40 OR #41 OR #42 OR #43 OR #44 OR #45

#47 #16 AND #31 AND #46 in Trials
